# Post Hoc Localization of Beam F3 Stimulation Targets: An MRI-Derived Geodesic Approach for Refined TMS E-Field Simulations

**DOI:** 10.64898/2026.06.21.26356164

**Authors:** Severin Schramm, Julia Ten Pas, Daniele Calabrò, Julia Jakubetz, Malte Szillat, Joan Koti, Minyan Huang, Su Hwan Kim, Michael Woletz, Jan Kirschke, Dennis M. Hedderich, Nico Sollmann, Martin Tik, Ulrike Vogelmann

**Affiliations:** Institute for Diagnostic and Interventional Neuroradiology, School of Medicine and Health, TUM Klinikum rechts der Isar, Technical University of Munich, Munich, Germany; German Center for Mental Health (DZPG), partner site München-Augsburg, Germany; Institute for Cardiovascular Radiology and Nuclear Medicine, German Heart Center Munich, School of Medicine and Health, Technical University of Munich, Munich, Germany; Clinic and Policlinic for Psychiatry and Psychotherapy, School of Medicine and Health, TUM Klinikum rechts der Isar, Technical University of Munich, Munich, Germany; MR Center of Excellence, Center for Medical Physics and Biomedical Engineering, Medical University of Vienna, Vienna, Austria; TUM-Neuroimaging Center, Klinikum rechts der Isar, Technical University of Munich, Munich, Germany; Department of Diagnostic and Interventional Radiology, University Hospital Ulm, Ulm, Germany; Department of Nuclear Medicine, University Hospital Ulm, Ulm, Germany

**Author notes:** **Address for correspondence:** Severin Schramm, Institute for Neuroradiology Tel.: 089/4140-7639, Klinikum rechts der Isar, Ismaninger Str. 22, 81675 Munich, Germany. **Complete Contact Information:** Severin Schramm, MD; Julia Ten Pas, cand. med.; Daniele Calabro, cand. med.; Julia Jakubetz, cand. med.; Malte Szillat, cand. med.; Su Hwan Kim, MD; Jan Kirschke, MD; Dennis Hedderich, MD, MBA; Nico Sollmann, MD, PhD, Institute for Neuroradiology.: 089/4140-4652, Klinikum rechts der Isar, Ismaninger Str. 22, 81675 Munich, Germany; Joan Koti, MD; Ulrike Vogelmann, MD, Clinic and Policlinic for Psychiatry and Psychotherapy.: 089/4140-4201, Klinikum rechts der Isar, Ismaninger Str. 22, 81675 Munich, Germany; Minyan Huang, MSc; Michael Woletz, PhD; Martin Tik, PhD; MR Center of Excellence, Center for Medical Physics and Biomedical Engineering, Medical University of Vienna, Spitalgasse 23, 1090 Vienna, Austria.

**Keywords:** Personalization, Therapy, Individualization, Transcranial Magnetic Stimulation, Neuronavigation

## Abstract

**Background:** Transcranial magnetic stimulation (TMS) targeting the left dorsolateral prefrontal cortex (dlPFC) is an established treatment option in major depressive disorder. One of the most common approaches for targeting the dlPFC is the Beam F3 method, which determines the stimulation site (F3_Beam_) as a function of external cranial measurements. Precise knowledge of the individual stimulation site is essential for imaging-based analyses of TMS effects. However, due to the method’s reliance on individual anatomy, retrospective identification of F3_Beam_ targets across cohorts is challenging, limiting the analysis of existing datasets. We developed a scalable method to reconstruct subject-specific F3_Beam_ target locations for e-field simulations based on structural imaging.

**Methods:** High-resolution three-dimensional (3D) T1-weighted MRI was used to generate individual scalp meshes via the “Simulation of Non-Invasive Brain Stimulation” (SimNIBS) software. Subject-specific anatomical distances and coordinates of interest were measured geodesically using a Python-based script to reconstruct the individual F3_Beam_ targets. Validation included a retrospective comparison between digital geodesic measurements and manual cranial measurements in 20 patients and a prospective comparison with MR-visible scalp markers in 2 healthy controls. To assess the impact of our targeting algorithm on e-field simulations, volumetric e-field maps based on three potential targets (F3_Beam,_ F3_MNI_, F3_Geo_) were generated in SimNIBS and compared using voxel-wise statistics in SPM12.

**Results:** Retrospective analysis revealed a systematic bias towards higher in vivo measurements compared to digital geodesic measurements, though deviations in the final distances determining F3_Beam_ (x_Beam_ and y_Beam_) were minimal (Δx_Beam_: 0.11 ± 0.08 cm; Δy_Beam_: 0.14 ± 0.21 cm). Prospective validation demonstrated that F3_Beam_ coordinates better matched in vivo coil positions than group-template-derived targets (F3_MNI_). Group-level analysis showed method-dependent clustering of coil positions with corresponding voxel-wise e-field differences.

**Conclusions:** Individualized geodesic measurements may enable accurate, scalable and retrospective identification of Beam F3 targets and coil orientations. This approach may yield more accurate e-field simulations than group-template based targeting and provides a practical method for retrospective analysis of existing TMS treatment cohorts. This could be leveraged to identify response predictors or imaging-based biomarkers of treatment response.

## 1. Introduction

Transcranial magnetic stimulation (TMS) is increasingly used in the therapy of a variety of psychiatric disorders ^1–9^. One of the most well-established indications for TMS treatment is major depressive disorder (MDD) ^1, 2, 6, 10^. The pathophysiological model underlying TMS usage presumes an imbalance between a hyperactive subgenual anterior cingulate cortex (sgACC) and hypoactive dorsolateral prefrontal cortex (dlPFC) in MDD patients ^5, 10–12^. This imbalance is adressed by stimulating the left dlPFC, which indirectly inhibits the sgACC ^5, 10–12^.

There are several ways to identify a dlPFC stimulation target on a subject’s head. While more elaborate methods incorporate functional MRI guidance and/or neuronavigation systems, one of the most common methods used is the Beam F3 method ^13, 14^. In this approach, three distances are measured directly on the patient’s head, i.e., tragus-tragus distance, nasion-inion distance, and head circumference. The stimulation site is then determined via two calculated target vectors x_Beam_ and y_Beam_ ^13, 14^. The method has seen widespread adoption due to its ease of application ^15–17^.

Recent advances in TMS research leverage patient-specific electric field (e-field) modeling to infer the spatial distribution of TMS-induced effects ^17–21^. By simulating how the brain’s geometry and structural properties shape the TMS-induced e-field, more precise estimations of stimulated regions are made possible. This has helped to predict TMS effects both in clinical populations and healthy controls ^17–21^. For example, individualized stimulation targets have been combined to inform both functional MRI and diffusion MRI analyses, enabling prediction of individual clinical outcomes in depression therapy ^17, 22^.

Despite enhancing imaging-based analyses, subject-specific Beam F3 localization and e-field simulations are still not widely implemented. This is likely due to the complexity of post hoc reconstruction of individual Beam F3 stimulation sites derived from multi-step manual measurements on the subjects’ head. While neuroimaging in combination with dedicated MR-visible markers can address this problem ^17^, this requires dedicated MRI acquisitions, ideally conducted prospectively. Additionally, using a group-template-derived F3 electrode coordinate may serve as an approximation, but can introduce discrepancies due to not reflecting the actual targeting procedure ^23^.

A robust and scalable method for reconstructing the individual Beam F3 stimulation site using structural imaging alone could unlock retrospective analyses of large, hitherto underexplored cohorts. Additionally, it could support treatment planning and patient-specific targeting through e-field informed individualization.

To address this unmet need, we developed a method for simulating Beam F3 target locations using patient-specific scalp models and individual geodesic measurements. We describe the retrospective validation of our method in a cohort of 20 patients and prospective validation in two healthy controls (HC).

## 2. Methods

### 2.1. Ethics

The imaging data utilized in this study were collected within an existing clinical study approved by the local ethics committee of the Technical University of Munich (project number: 2024-220_3-S-CB). Written informed consent for scientific usage of the acquired imaging was obtained from each study participant prior to study inclusion.

### 2.2. Imaging

Imaging was performed on a 3T scanner (Ingenia; Philips Medical Systems, The Netherlands B.V.) using a 32-channel head coil. Both MDD patients as well as HC underwent a sequence protocol which included a high-resolution three-dimensional (3D) T1-weighted sequence (repetition time [TR]/echo time [TE]: 9/4 ms, 1 mm^3^ isovoxel covering the whole head) for structural imaging. Patients furthermore underwent a T2-like 3D magnetization transfer (MT)-weighted sequence (TR = 48 ms, α = 6°, TE = 7 ms, MT saturation flip angle αMT = 220°, MT pulse duration tMT = 8 ms) as part of a larger scanning protocol, which was used to supplement cortical surface reconstructions.

### 2.3. Geodesic Measurements

The goal of the project was to create a digital step-by-step reproduction of the Beam F3 method workflow. The desired output was the determination of the F3_Beam_ target and a corresponding reference direction coordinate (Ref _Beam_, indicating coil direction) for e-field simulations.

For a given subject, F3_Beam_ is found as a combination of two coordinate vectors x_Beam_ and y_Beam_, which in turn are functions of individual anatomical distances ^13^.

Ref _Beam_ constitutes a point on the midline scalp which, when connected with F3_Beam_ forms a 45° intersection with the midline ^13^.

#### 2.3.1. Scalp Model Generation

We used the SimNIBS 4.5 charm pipeline ^24^ to generate individual scalp meshes from structural imaging. Cortical reconstruction was using Freesurfer-based surface segmentation (version 8.0.0) ^25^. In patients, the charm pipeline was supplemented by the above mentioned T2-like imaging.

#### 2.3.2. Geodesic Distance Measurements

The individual scalp meshes were subsequently used for geodesic measurements. The soft tissue component of the meshes was extracted from the total mesh and any mesh defects were corrected using PyMeshFix (version 0.17.1) ^26^ to create a watertight mesh.

Geodesic distances between points of interest were computed using the PyGeodesic library (version 0.1.9) ^27^. As priors, we extracted the coordinates of the following points from the SimNIBS m2mfolder: nasion (Nz), inion (Iz), left tragus (lTr), right tragus (rTr), vertex electrode (Cz), electrode positions T7, T8, and Fpz according to the SimNIBS coordinates for the 10-10 electroencephalography (EEG) system ^28^.

For comparison with the geodesic measurements, we also exported the F3 electrode position (F3_MNI_) as output from the SimNIBS charm pipeline according to the 10-10 EEG system obtained via reverse-transformation from MNI152 space ^28, 29^. The accuracy of the key anatomical landmarks (Nz, Iz, lTr and rTr) was confirmed visually by inspecting the SimNIBS head mesh for each subject. Additionally, we defined a fiducial point “EB” at one third of the geodesic path of Nz-Fpz to approximate the eyebrow level for head circumference measurement ^13^.

To reconstruct the manual Beam F3 targeting procedure performed on the individuals’ head, we measured the distances Nz-Cz-Iz, lTr-Cz-rTr, and EB-T7-Iz-T8-EB (head circumference) on the individual scalp mesh and exported these distances to a .csv file (Figure 1B).

**Figure 1:**
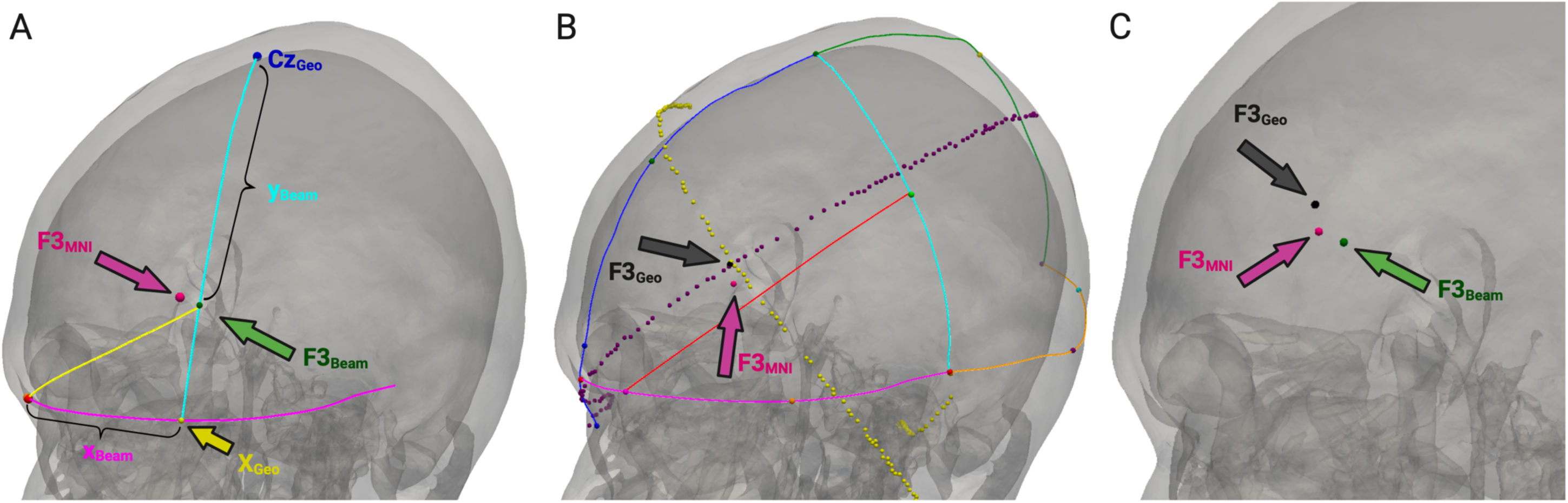
Exemplary Geodesic Measurements. demonstrates example screenshots from the geodesic measurement process. Depicted points include F3_Beam_ (Beam F3 target determined by our geodesic approach, green), F3_MNI_ (F3 electrode position determined by reverse transformation from MNI152 group space, magenta) and F3_Geo_ (F3 electrode position determined by geodesic approach, black). A) Determination of F3_Beam_. After calculating x_Beam_ and y_Beam_ distances, F3_Beam_ (green, green arrow) is determined by going the distance x_Beam_ along the head circumference path to reach X_Beam_ (yellow dot) and subsequently going the distance y_Beam_ from Cz towards X_Beam_. B) Determination of F3_Geo_. F3_Geo_ (black arrow) is determined by calculating the intersection of equidistance lines between F7 and Fz (violet dots) as well as FP1 and C3 (yellow dots). A magenta dot (magenta arrow) indicates the position of F3_MNI_, the MNI coordinate for F3 reverse-transformed into subject space. C) Exemplary comparison of F3_Geo_ (black), F3_MNI_ (magenta) and F3_Beam_ (green).

#### 2.3.3. Beam F3 Target Localization

We used the online tool provided in the initial publication of the Beam F3 method to input the anatomical distances for nasion-inion, tragus-tragus and head circumference (i.e., Nz-Cz-Iz, lTr-Cz-rTr, and EB-T7-Iz-T8-EB) and generate the x_Beam_ and y_Beam_ distances ^13^. Specifically, we used the adjusted y_Beam_ distance (adding 0.35 cm) as established by Mir-Moghtadaei et al. ^14^. The resulting x_Beam_ and y_Beam_ distances were then passed to our geodesic-based script to finalize the target determination according to the following steps: We calculated the geodesic distance of Nz-Cz-Iz and defined the midpoint (M) on this path. From M, we calculated the distance lTr-M-rTr and similarly identified the midway point on this path. This point was defined as the geodesic vertex electrode position (Cz_Geo_). Importantly, Cz_Geo_ is approximately equidistant to both Nz and Iz as well as to lTr and rTr, marking a key reference point in the Beam F3 targeting procedure ^13^. Next, we identified the point X_Geo_ by measuring the distance x_Beam_ along the path EB-T7, following the same path used in the head circumference measurement. Lastly, we identified the target F3_Beam_ by measuring the distance y_Beam_ along the path Cz_Geo_-X_Geo_ (Figure 1A). The coordinates were exported to a .csv file for further analysis.

#### 2.3.4. Individual Geodesic Determination of F3 Electrode Position (F3_Geo_)

As an additional reference for comparisons, we performed an individual geodesic determination of the F3 electrode position (F3_Geo_) according to the 10-10 EEG system. This yields a second, independent F3 coordinate based on the standard 10-10 layout rather than the Beam F3 method.

For this, we used the SimNIBS points Nz, Iz, lTr, rTr, and T7, as well as the previously calculated Cz_Geo_ to construct the electrode positions T3, Fz, C3, Fp1, and F7 based on geodesic measurements.

Specifically, Fz was determined at 30 % of the Nz- Cz_Geo_-Iz distance starting from Nz; T3 was defined as the midpoint of the EB-T7-Iz path; C3 was defined as the midpoint of the Cz_Geo_-T3 path; Fp1 was located at 10% of the EB-T7-Iz distance, starting from EB; F7 was located 30% of the EB-T7-Iz distance starting at EB.

We then defined F3_Geo_ as the point equidistant to Fz and F7 as well as equidistant to C3 and Fp1 (Figure 1B).

#### 2.3.5. Reference Direction Definition

In the Beam F3 method, coil orientation is determined by holding the coil handle in a 45° angle pointing away from the midline ^13^. In clinical practice, this is achieved by using a flexible measurement tool to define the geodesic line which connects the coil center with the midline at a 45° angle.

To replicate this process, we determined the coil reference direction (i.e., the direction the coil is pointing towards) by identifying the point on the path Nz-Cz_Geo_ which, when connected to F3_Beam_, most closely results in a 45° intersection. This point was named Ref_Beam_ and its coordinates were exported to a .csv file. Analogous points were identified and exported by looking for a 45° intersection when connecting to the coordinates of F3_MNI_ as well as F3_Geo_.

#### 2.3.6. Validation of Measurements

To validate our measurements, we employed a retrospective and a prospective comparison.

Retrospectively, we collected in-vivo scalp measurements (Nz_viv_-Cz_viv_-Iz_viv_, lTr_viv_-Cz_viv_-rTr_viv_, Circumference_viv_, X_viv_, and Y_viv_) from 20 patients who had undergone depression treatment in our local psychiatry department. Head models were visually inspected to ensure absence of severe motion- or segmentation-related artifacts. We then compared these manually measured to our image-based measurements using Bland-Altman plots, concurrence correlation coefficient (CCC) analysis ^30^, and Passing-Bablok regression ^31^. Statistical analysis was implemented in RStudio (version 2025.05.1+513) using R (version 4.5.1).

Prospectively, we performed an MRI- based validation in 2 HC. Each participant underwent initial structural imaging followed by manual head measurements performed by a psychiatrist (with one year of experience in the Beam F3 targeting approach). The measured coil center and coil direction were marked using MRI-visible capsules and a second MRI scan was acquired. Points of interest (F3_Beam_, F3_MNI_ and their respective reference direction coordinates) were calculated based on the initial imaging and then overlaid on the coregistered marker-enhanced imaging to visualize concordance between digitally measured points and the physically marked positions.

#### 2.3.7. Electric Field Bias Mapping

E-fields were simulated in SimNIBS 4.5 using the MagMore PMD70 coil model ^32^. The maximum current gradient was set to 1,00 x 1e6 A/s. For each patient, we simulated the three calculated coil positions (F3_MNI_, F3_Geo_, and F3_Beam_). The resulting e-fields were exported as .nifti files in both subject space and Montreal Neurological Institute (MNI)-152 space for group level analysis. Additionally, coil positions for each subject were transformed to MNI152 space and overlaid on a template to visualize clustering of coil positions across targeting methods.

We used SPM12 ^33^ to implement a flexible factorial statistical test design for voxel-wise comparisons of the generated e-fields. Specifically, we modeled the e-field magnitude as the dependent variable and the targeting method (F3_Default_, F3_Geo_, Beam F3) as a nominal factor. Analyses were performed using an explicit binary mask for the left dlPFC as previously employed by Cash et al. ^3^, given that e-field magnitude rapidly decreases with distance from the coil location. We generated t-contrast maps for the differences between targeting methods. A statistical threshold of p = 0.001 (voxel-level) and p = 0.05 (family-wise error-corrected, cluster level) was applied.

## 3. Results

### 3.1. Retrospective Validation

Across all measured distances, in vivo values were systematically higher than digital measurements digitally (Figure 2). Mean differences (in vivo minus digital) between measured distances were 1.23 ± 1.00 cm for Nz-Cz-Iz; 2.72 ± 0.91cm for lTr-Cz-rTR; 1.08 ± 0.66 cm for head circumference; 0.11 ± 0.08 cm for x_Beam_ and 0.14 ± 0.21 cm for y_Beam_. Table 1 summarizes the measurements and differences between digital- and in vivo values.

**Figure 2:**
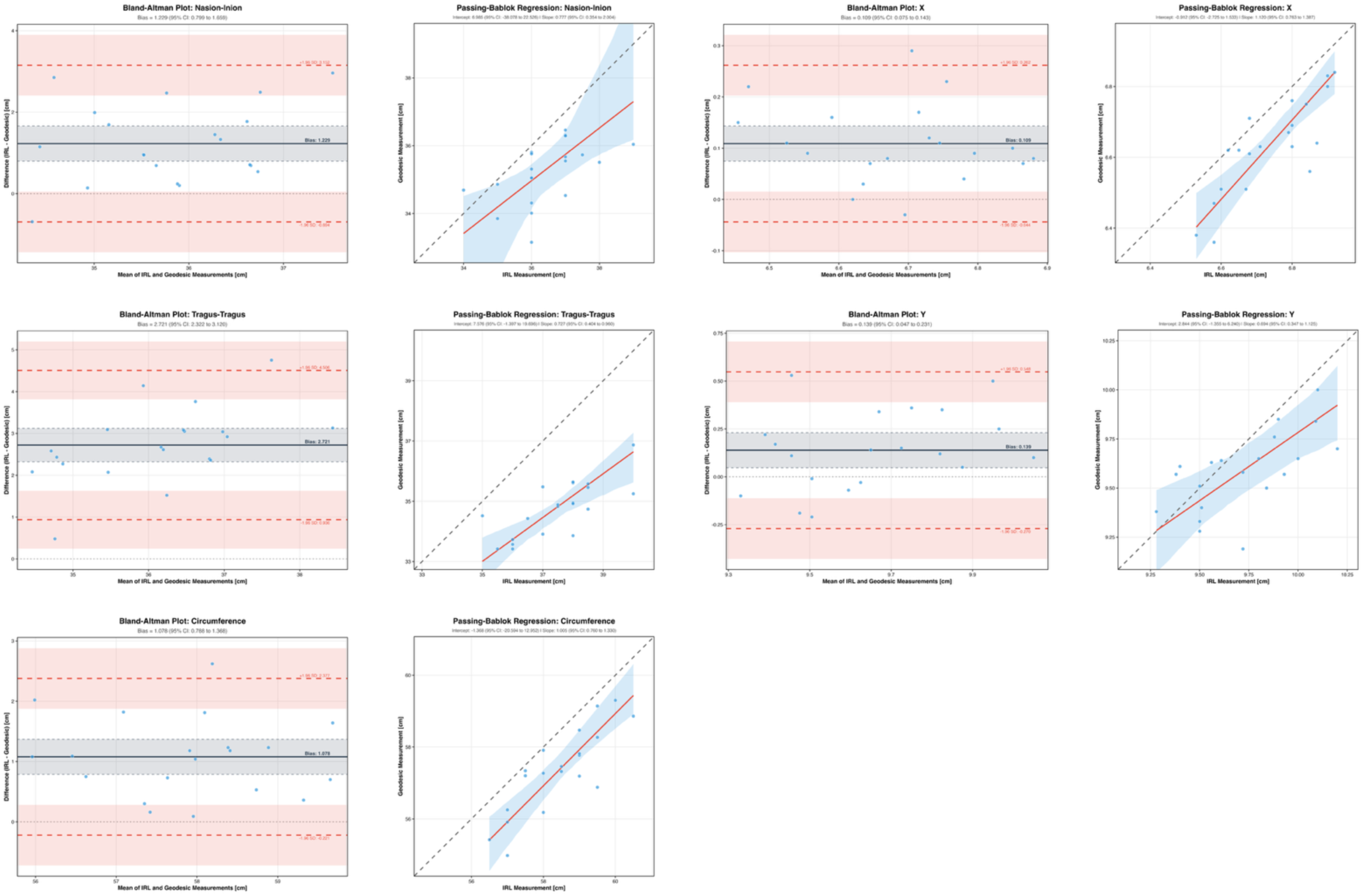
Bland-Altman Plots and Passing-Bablok Regressions for Geodesic and In Vivo Measurements. Analyses reveal a systematic bias towards higher in vivo values across all anatomical distances. Despite this, scaling between in vivo and digital values was acceptable within the tested range.

**Table 1:** Overview of Distances Measured in Vivo and Digitally. summarizes the distances measured in vivo and their corresponding digital geodesic measurements. Abbreviations: standard deviation (SD).

| **Distance** | **In Vivo Measurement;**<br>**Mean ± SD [cm]** | **Digital Measurement;**<br>**Mean ± SD [cm]** | **Delta of Measurements;**<br>**Mean ± SD [cm]** |
| --- | --- | --- | --- |
| **Nasion-Inion** | 36.43 ± 1.12 | 35.20 ± 0.90 | 1.23 ± 1.00 |
| **Tragus-Tragus** | 37.48 ± 1.36 | 34.75 ± 0.91 | 2.72 ± 0.91 |
| **Circumference** | 58.43 ± 1.12 | 57.35 ± 1.2 | 1.08 ± 0.66 |
| **X<sub>Beam</sub>** | 6.74 ± 0.12 | 6.63 ± 0.14 | 0.11 ± 0.08 |
| **Y<sub>Beam</sub>** | 9.72 ± 0.27 | 9.58 ± 0.20 | 0.14 ± 0.21 |

The CCC analysis indicated suboptimal agreement (< 0.9 for all distance types) between in vivo and geodesic measurements (Table 2). Passing-Bablok regression indicated acceptable scaling overall (Figure 2; Table 3) with the potential exception of lTr-Cz-rTR (95 % confidence interval for slope: 0.4-0.96, excluding 1).

**Table 2:** Concordance Correlation Coefficient Analysis. summarizes the metrics from the Concordance Correlation Coefficient (CCC) analysis comparing the in vivo distances with their corresponding digital geodesic measurements. Abbreviations: Concordance Correlation Coefficient (CCC), confidence interval (CI).

| **Distance** | **CCC Estimate and 95% CI** | **Pearson Correlation Coefficient** |
| --- | --- | --- |
| **Nasion-Inion** | 0.30 [0.04; 0.52] | 0.54 |
| **Tragus-Tragus** | 0.18 [0.06; 0.29] | 0.75 |
| **Circumference** | 0.57 [0.32; 0.74] | 0.83 |
| **X<sub>Beam</sub>** | 0.60 [0.34; 0.77] | 0.82 |
| **Y<sub>Beam</sub>** | 0.52 [0.19; 0.74] | 0.63 |

**Table 3:** Passing-Bablok Regression. summarizes the metrics from the Passing-Bablok regression comparing the in vivo distances with their corresponding digital geodesic measurements. In the Passing-Bablok regression, an intercept ≠ 0 indicates a systematic bias, while the slope indicates whether scaling is comparable across the two measurement methods with 1 indicating perfect scaling. Abbreviations: Confidence interval (CI).

| **Distance** | **Intercept and 95% CI** | **Slope and 95% CI** |
| --- | --- | --- |
| **Nasion-Inion** | 6.99 [-38.08; 22.53] | 0.78 [0.35; 2.00] |
| **Tragus-Tragus** | 7.58 [-1.40; 19.70] | 0.73 [0.40; 0.96] |
| **Circumference** | -1.37 [-20.59; 12.95] | 1.00 [0.76; 1.33] |
| **X<sub>Beam</sub>** | -0.91 [-2.73; 1.53] | 1.12 [0.76; 1.39] |
| **Y<sub>Beam</sub>** | 2.84 [-1.36; 6.24] | 0.69[0.35; 1.13] |

### 3.2. Prospective Validation

#F3_Beam_ demonstrated better overlap with the in vivo marked points compared to F3_MNI_. In both individuals, F3_Beam_ was located within the marker structure, while F3_MNI_ remained more anterior. Coil direction as indicated by Ref_Beam_, was highly comparable to the reference direction measured in vivo (Figure 3).

**Figure 3:**
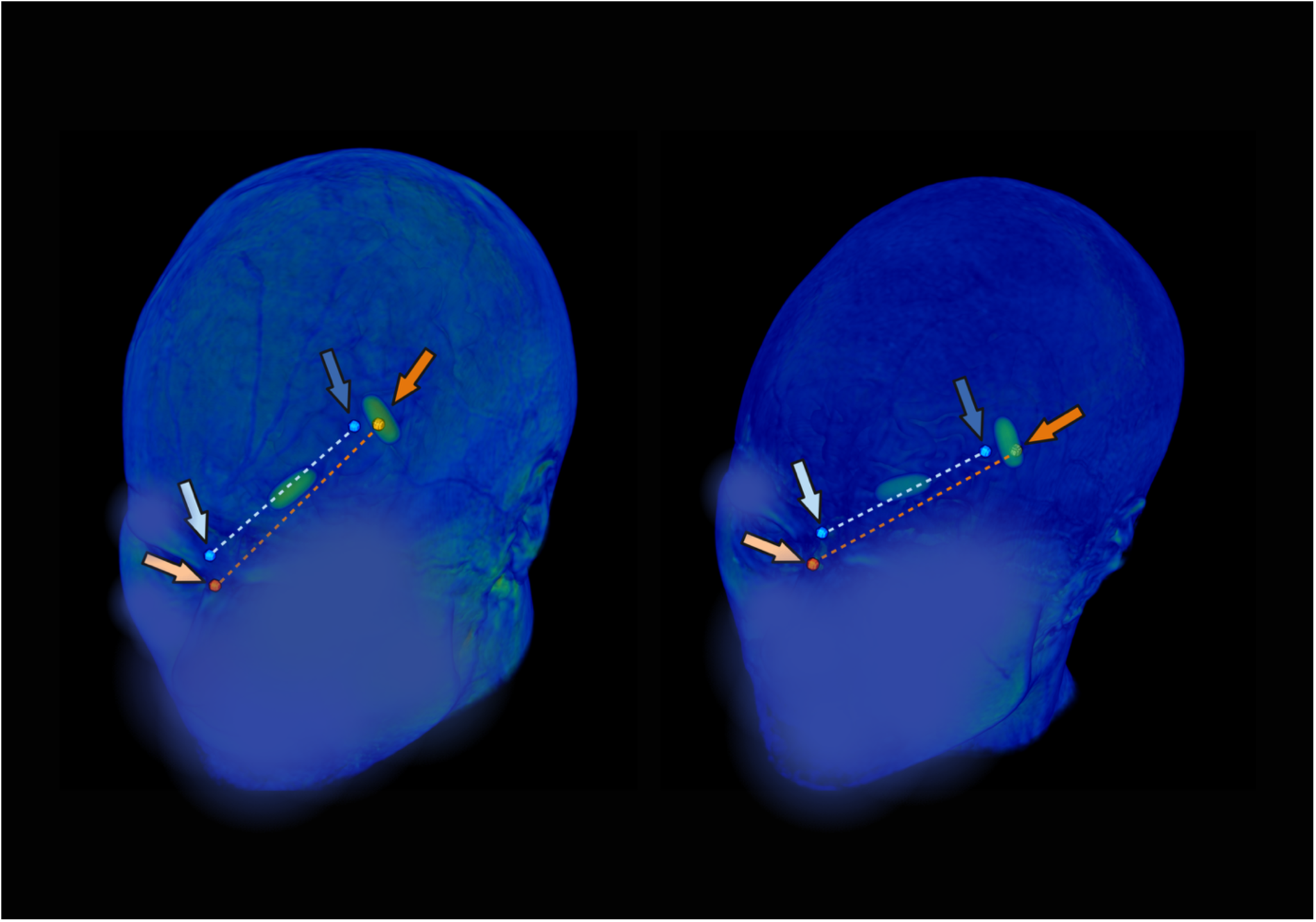
Prospective Targeting Validation. Scalp reconstructions with overlayed target coordinates. MRI markers on the scalp in green indicate the physically marked positions for coil center and reference direction. Blue points and arrows indicate F3_MNI_ and its corresponding reference target for 45° coil orientation (blue dotted line); orange points and arrows indicate F3_Beam_ and Ref_Beam_ points. In both subjects, F3_Beam_ lies within the marker structure while F3_MNI_ is located anterior to the marker. Of note, the dotted lines are overlaid projections and do not reflect the true 3D path on the head surface used in the reference point calculations.

### 3.3. Electric-Field Bias Mapping

Voxel-wise comparisons of e-fields based on F3_MNI_, F3_Geo_, and F3_Beam_ indicated significant differences in simulated e-field distributions, corresponding to the observed clustering of coil positions (Figure 4). While F3_MNI_ naturally converged to a single MNI space coordinate at the group level, F3_Geo_ clusters were located more posteriorly, and F3_Beam_ positions more posterior-inferiorly. Consequently, F3_MNI_ resulted in systematically higher e-fields in the superior-anterior dlPFC, and lower e-fields them in the posterior-inferior dlPFC compared to F3_Beam_ (Figure 4). A medial-lateral bias was observed when comparing F3_Geo_ derived e-fields to F3_Beam_ e-fields (Figure 4).

**Figure 4:**
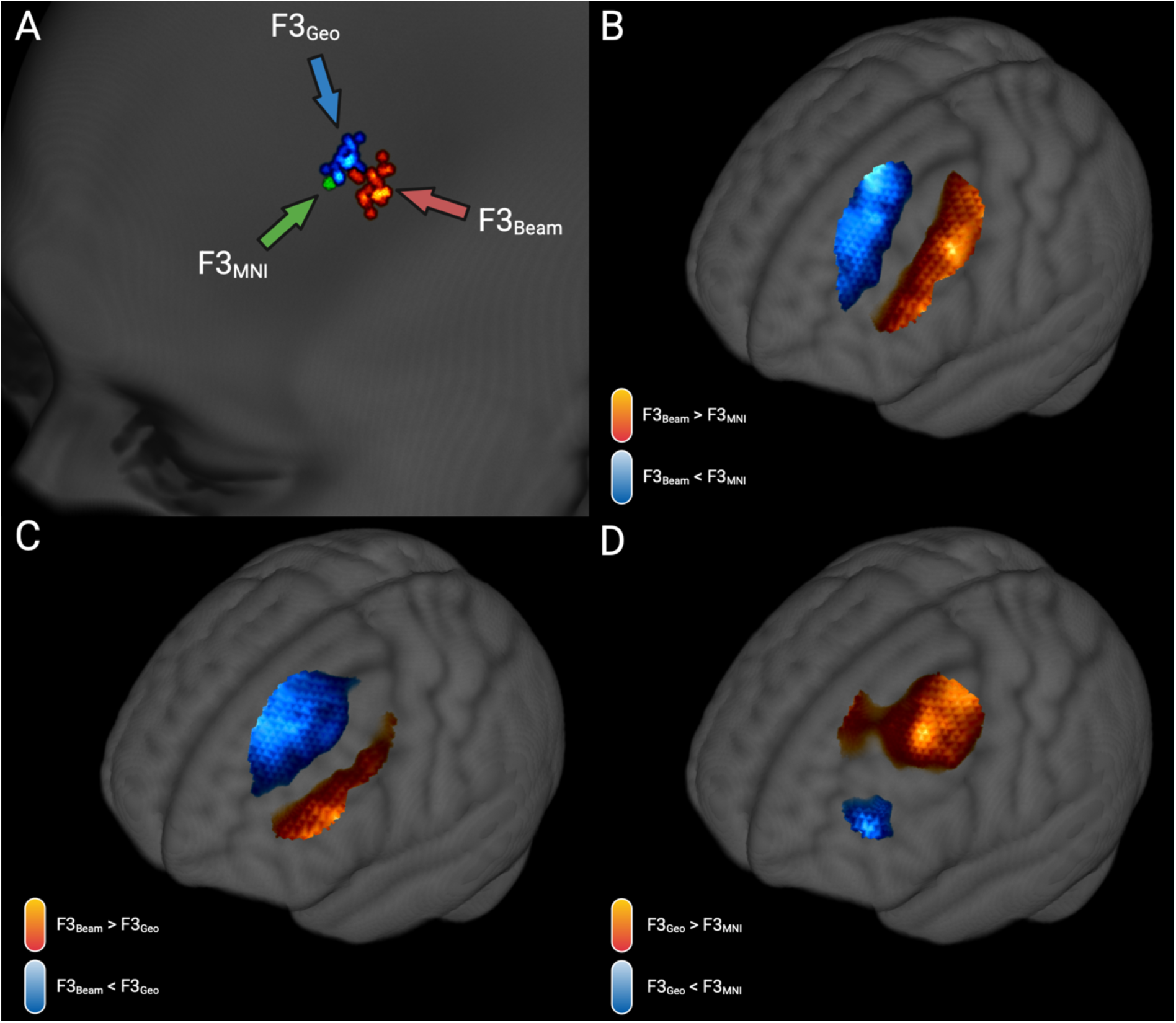
Target Locations at the Group Level and E-Field Biases. A) Targets positions by method overlaid onto an MNI152 template scalp. The green point (green arrow) indicates F3_MNI_, which converges to a single MNI152 coordinate across subjects. Red points (red arrow) indicate F3_Beam_ coordinates, clustering more posterior and inferior compared to F3_MNI_. Blue points (blue arrow) indicate coordinates for F3_Geo_, clustering slightly more posterior compared to F3_MNI_. B) Thresholded T-contrast for E-fields of F3_Beam_ vs. F3_MNI_. The warm-colored cluster indicates areas with higher e-field magnitude for F3_Beam_, while the cool-colored cluster indicates areas with lower e-field magnitude for F3_Beam_. A threshold of voxel-level p < 0.001, cluster-level p < 0.05 FWE-corrected was applied. C) Thresholded T-contrast for E-fields of F3_Beam_ vs. F3_Geo_. The warm-colored cluster indicates areas with higher e-field magnitude for F3_Beam_, while the cool-colored cluster indicates areas with lower e-field magnitude for F3_Beam_. A threshold of voxel-level p < 0.001, cluster-level p < 0.05 FWE-corrected was applied. D) Thresholded T-contrast for E-fields of F3_Geo_ vs. F3_MNI_. The warm-colored cluster indicates areas with higher e-field magnitude for F3_MNI_, while the cool-colored cluster indicates areas with lower e-field magnitude for F3_MNI_. A threshold of voxel-level p < 0.001, cluster-level p < 0.05 FWE-corrected was applied.

## 4. Discussion

We present a method for retrospective identification of Beam F3 TMS targets based on individualized geodesic scalp measurements. Our results indicate that this individualized method provides more accurate coil positions than group-level MNI-based targeting of the F3 electrode position (Figure 3), and that group-level assumptions may introduce systemic biases in calculated e-fields (Figure 4).

### 4.1. Measurement Validity

Our retrospective comparison of digitally measured values and in vivo values yielded mixed results. The CCC analysis suggested poor reliability, which we interpret as largely reflecting a systematic upward bias in the in vivo measurements. However, inspection of the Passing-Bablok regression plots indicated that geodesic distances were well scaled across the observed ranges despite an overall shift towards lower digital values.

We suspect that the higher in vivo values arise from how the measurement procedure is implemented in vivo. Our mesh-based approach measures directly on the scalp, whereas in vivo measurements are usually performed with subjects wearing a textile cap that compresses hair and adds an additional layer on top of the scalp. This likely results in higher distances across all measurements. Importantly, for the Beam F3-relevant distances x_Beam_ and y_Beam_, deviations from the in vivo values were minimal and remained in the low millimeter range (Table 1).

Furthermore, prospective validation direct distance comparisons showed that our approach approximates the Beam F3 targeting procedure more accurately than MNI-based targeting of the F3 electrode position (Figure 3). Additionally, our process is fully deterministic and can be tailored towards other targets such as alternative EEG coordinates.

### 4.2. Use Cases

Although neuronavigation of TMS is becoming increasingly common, non-navigated application of stimulation remains widespread globally due to its lower cost and ease of application. In this context, an accurate way to retrospectively simulate the individual e-field applied during therapy could unlock large cohorts for add-on analyses, enabling more sophisticated investigations of potential response predictors and imaging-based biomarkers of treatment response such as functional or structural connectivity patterns specific to the individually stimulated cortical areas. Importantly, e-field intensity is assumed to influence clinical outcome ^34, 35^. Therefore, precise e-field modeling is crucial, as the strength of the induced e-field at the intended target, typically the dlPFC, may directly affect clinical outcomes. Inaccurate localization of the stimulation site can lead to substantial deviations in the modeled e-field intensity at the target region, thereby confounding the interpretation of dose–response relationships. Consequently, improving the precision of e-field estimation at the true stimulation site is essential for reliably assessing and potentially optimizing the clinical efficacy of TMS interventions.

A prior study had already indicated discrepancies between the Beam F3 target and the F3 electrode spot according to the 10-20 EEG system ^23^. Our results similarly show that F3_Beam_ coordinates cluster distinctly from both F3_MNI_ and F3_Geo_ coordinates, with corresponding biases in the simulated e-field (Figure 4). One of the advantages of using personalized measurements is exemplified here: target locations cluster around a given spot yet vary more across individuals than can be captured in a single MNI coordinate (Figure 4). Previous work has already demonstrated the predictive power of individualized e-field mappings in various contexts ^17, 18, 36, 37^. However, these approaches have hitherto relied on either neuronavigation or marker-based recordings of stimulation sites ^17, 18, 36, 37^.

### 4.3. Limitations

Physical measurements are usually performed on the patient’s head, often over a cap, rather than directly on the scalp, which may introduce a bias in our method regarding the measurements of Nz_viv_-Cz_viv_-Iz_viv_, lTr_viv_-Cz_viv_-rTr_viv_ and Circumference_viv_. Nonetheless, our results suggest a meaningful improvement over non-individualized, MNI-based targeting, better reflecting the actual stimulation sites and thus generating more accurate, individualized e-field maps.

Second, our approach relies on high-quality structural imaging, including at least a high-resolution 3D T1-weighted sequence and appropriate segmentation of mesh components in the SimNIBS charm pipeline. Although we cannot provide e-fields for patients with no imaging data at all, recent advances in the field of synthetic MRI ^38^ could potentially generate individual high-resolution T1-weighted imaging for patient with lower resolution data, and thus provide an avenue for geodesic Beam F3 targeting reconstruction.

Third, our approach relies on multiple starting coordinates (e.g., Nz, Iz, lTr, or rTr), which are by default extracted from SimNIBS and thus are reverse-transformed from MNI152 space. This implementation allows efficient batch processing but is dependent on accurate transformations between spaces. Manual identification of these above-mentioned reference coordinates could potentially improve subject-specific accuracy but would come at the cost of added time required to visually identify the respective landmarks.

## 5. Conclusion

We present a method for accurate and individualized identification of the Beam F3 target and coil orientation based on structural imaging. This may allow for retrospective simulation of applied e-fields in TMS applications, such as for the treatment of depression. Our results indicate that this method yields more accurate e-field simulations than MNI-based targeting, and results in systematically different e-fields. This method can be employed to retrospectively investigate cohorts regarding potential response predictors or therapy-associated imaging markers for TMS.

## Data Availability

A research prototype of the code presented in this manuscript is available under "https://github.com/SeSchramm/GeoBeam.git". The available code covers the core functionalities presented in the script but is still undergoing active development.
Other data used in this manuscript is available upon reasonable request to the authors.

## 6. Acknowledgements

None.

## 7. Disclosures

-

## Notes

### Competing Interest Statement

The authors have declared no competing interest.

### Author Declarations

The Ethics committee of University Hospital Rechts der Isar gave ethical approval for this work.

